# Integrative Mendelian Randomization for Detecting Exposure-by-group Interactions Using Group-Specific and Combined Summary Statistics

**DOI:** 10.1101/2025.01.26.25321136

**Authors:** Ke Xu, Nathaniel Maydanchik, Bowei Kang, Jianhai Chen, Qixiang Chen, Gongyao Xu, Shinya Tasaki, David A. Bennett, Lin S. Chen

## Abstract

Interactions between risk factors and covariate-defined groups are commonly observed in complex diseases. Existing methods for detecting interactions typically require individual-level data. The data availability and the measurements of risk exposures and covariates often limit the power and applicability in assessing interactions. To address these limitations, we propose int2MR, an integrative Mendelian randomization (MR) method that leverages GWAS summary statistics on exposure traits and group-separated and/or combined GWAS statistics on outcome traits. int2MR can assess a broad range of risk exposure effects on diseases and traits, revealing interactions unattainable with incomplete or limited individual-level data. Simulation studies demonstrated that int2MR effectively controls type I error rates under various settings while achieving considerable power gains with the integration of additional group-combined GWAS data. We applied int2MR to two data analyses. First, we identified risk exposures with sex-interaction effects on ADHD, and our results suggested potentially elevated inflammation in males. Second, we detected age-group-specific risk factors for Alzheimer’s disease pathologies in the oldest-old (age 95+), many of which were related to immune and inflammatory processes. Our findings suggest that reduced chronic inflammation may underlie the distinct pathological mechanisms observed in this age group. int2MR is a robust and flexible tool for assessing group-specific or interaction effects, providing insights into disease mechanisms.

## Introduction

Complex diseases often arise from a combination of genetic, environmental, and biological factors, resulting in varied effects of risk factors across different subgroups., ^1^,^2^These group-specific risk effects, also known as exposure-by-group interaction effects, play a critical role in disease mechanisms.^3^,^4^ For example, genetic variations may influence susceptibility to risk factors such as diet, pollution, or lifestyle within specific populations.^5–11^ Biological differences, such as age and sex, may lead to different disease pathways across groups.^12–21^ Additionally, social determinants of health, including access to healthcare and socioeconomic status, may shape exposure-disease relationships differently for various groups.^22 –24^ Understanding these exposure-by-group interaction effects is crucial for advancing research on disease mechanisms and precision medicine. By considering group-specific effects, precision medicine can better address the unique risks of diverse populations and improve health outcomes, especially for vulnerable populations.

To assess exposure-by-group interaction effects, a common approach is interaction analysis, which estimates both the effects of risk exposures and their interactions with groups or covariates on disease outcomes.^25^ These analyses, though, are often constrained by small sample sizes and the limited availability of risk exposure and covariate measurements within individual studies. Moreover, unmeasured confounding variables can bias the results, complicating causal interpretations. Two-stage least squares (2SLS) methods have been applied to detect interactions while allowing for unmeasured confounding under certain assumptions.^26^ However, their applicability is also restricted to studies with individual-level data, and the power is constrained by the strength of the instruments in the data. Mendelian randomization (MR) is a powerful tool to evaluate the causal effects of risk exposures on complex disease outcomes, treating genetic variants associated with exposures of interest as instrumental variables (IVs).^27 –30^ Two-sample MR, which uses two sets of genome-wide association study (GWAS) summary statistics as input, has achieved many successes in assessing the causal effects of complex traits as exposures on various diseases as outcomes. ^31–42^

Most existing MR methods are designed to assess total effects, with limited focus on detecting exposure-by-group interaction effects. While some recent MR methods were proposed to detect interaction effects,^3,4^ they require individual-level data on exposure, outcome, and the covariate group variables. The growing availability of GWAS summary statistics highlights the need for two-sample MR methods to detect interactions across covariate-defined groups using summary data.

Here we propose an *int*egrative MR method for detecting *int*eraction effects (int2MR) between exposures and covariate groups on complex diseases, using only summary statistics as input. The int2MR method integrates group-specific and group-combined GWAS summary statistics from multiple studies and consortia, enhancing the power to detect group-specific effects of risk exposures. Through extensive simulations and method comparisons, we demonstrated the advantages of the proposed int2MR method for detecting interactions and main effects. We applied the proposed int2MR method to two data analyses. In the first analysis, by integrating sex-stratified and sex-combined ADHD GWAS summary statistics from the Psychiatric Genomics Consortium ^43,44^ and other major consortia, we boosted the power for identifying exposures with sexinteraction effects on ADHD. In the second analysis, motivated by the observation that AD pathology peaks around age 95 and then declines, we sought to identify risk factors with age-group-specific effects on AD pathology in the oldest-old (defined as death age 95+) compared to the rest of the population. Using int2MR, we integrated age-groupstratified GWAS summary statistics from ROSMAP (Religious Orders Study and the Rush Memory and Aging Project) ^45^ with publicly available GWAS summary statistics on a wide range of risk exposures from major consortia. We identified multiple immune and inflammation-related exposures with age group-differential effects on AD pathology in the oldest-old, suggesting reduced inflammation and potentially distinct neuroinflammatory mechanisms in the oldest-old compared with the rest of the population. These analyses demonstrate int2MR’s flexibility in assessing exposure-by-group interactions across diverse traits and groups (e.g., socioeconomic or environmental factors) using GWAS summary statistics. It enhances detection power for main and interaction effects by integrating group-stratified and group-combined GWAS data.

## Methods

We propose the int2MR model to detect exposure-by-group interaction effects. We introduce two variations of the model, as illustrated in Figure 1. In this section, we will describe the input statistics and then present the model formulation and the estimation algorithms for both variations. Furthermore, the model is flexible and can be extended to integrate IV-to-outcome statistics from mixed groups with varying group compositions (see Supplemental Information for details).

**Figure 1:**
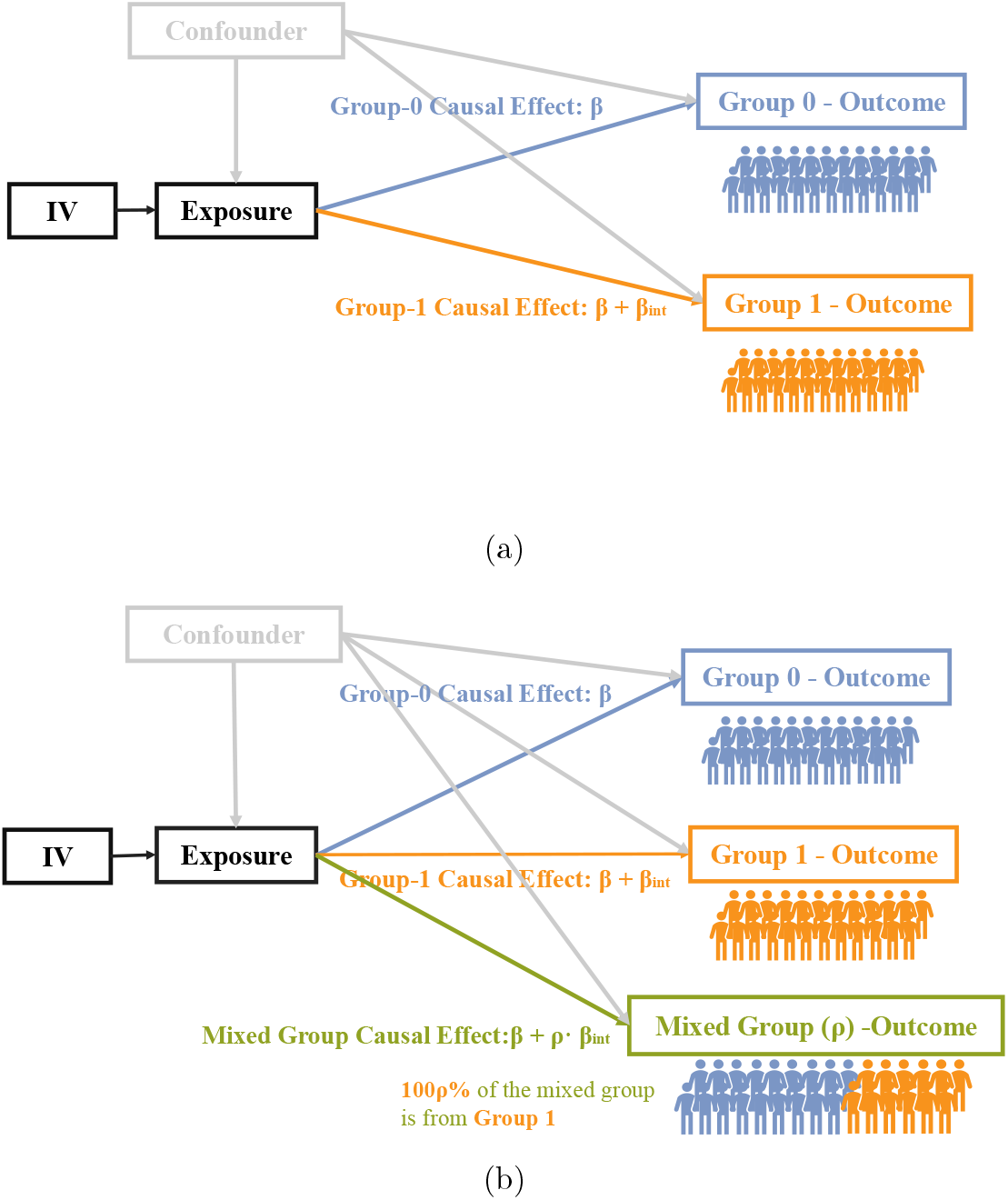
Illustrations of the proposed int2MR framework for detecting interaction effects. (a) The causal diagram of the int2MR model, using summary statistics from two group-specific IV-to-outcome GWASs as input. The model estimates the causal effect *β* for the reference group (Group 0) and *β* +*β*_int_ for the comparison group (Group 1), capturing group-specific effects. (b) int2MR using two sets of group-specific IV-to-outcome GWAS statistics and one set of group-combined GWAS statistics for input. The parameter *ρ* represents the proportion of samples from the comparison group in the mixed-group study. The IV-to-exposure GWAS statistics in int2MR analyses are all from standard group-combined GWAS analyses.

### int2MR: An integrative MR for detecting exposure-by-group interaction effects with group-seperated GWAS summary statistics

As illustrated in Figure 1(a), the first variation of int2MR takes the following summary statistics as input:

- IV-to-exposure statistics: Standard GWAS summary statistics for the exposure trait.
- Group-specific IV-to-outcome statistics: GWAS summary statistics for the outcome stratified by groups (i.e. reference group and comparison group).

For the *j*-th SNP (*j* = 1, 2, · · ·, *p*), let 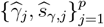 denote the observed marginal IV-to-exposure effect and its standard error obtained from a GWAS study for the exposure trait. Similarly, let 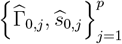 and 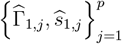 denote the observed marginal IV-to-outcome effects and their standard errors for the reference group (denoted by subscript 0) and the comparison group (denoted by subscript 1), respectively. The IV-to-outcome statistics for the two groups are obtained from group-stratified GWAS studies on the outcome trait. Suppose that 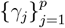 are the true IV-to-exposure effects, and 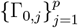 and 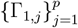 are the true IV-to-outcome effects for the reference and comparison groups, respectively. For the *j*-th SNP as an IV, we jointly model the IV-to-exposure effect and the IV-to-outcome effects in the reference and comparison groups as:

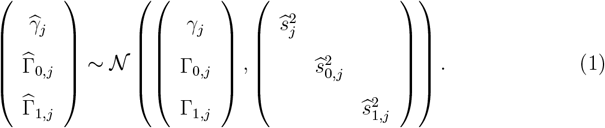

The true IV-to-outcome effects in the two groups are:

Reference group (Group 0) : Γ_*j*_ = *β* · *γ*_*j*_ + *α*_0,*j*_,

Comparison group (Group 1) : Γ_*j*_ = (*β* + *β*_int_) · *γ*_*j*_ + *α*_1,*j*_.

Here, 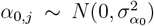 and 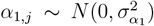 represent uncorrelated pleiotropic effects^37^ in the reference group and comparison group, respectively. The main effect *β* captures the causal effect in the reference group, while *β*_int_ represents the differential effect between the comparison and reference groups, i.e., the interaction effects. This framework allows for estimating and testing any linear combination of *β* and *β*_int_. For example, we can separately test for non-zero group-specific causal effects, *β* and *β* + *β*_int_, for the reference and comparison groups, respectively. Additionally, we can test for non-zero interaction effects, *β*_int_. Note that the off-diagonal elements in the covariance matrices of Model (1) are non-zero if there are sample overlaps among different GWAS studies.

### Flexible int2MR allowing the integration of group-specific and group-combined GWAS statistics

The proposed int2MR model can integrate group-specific and group-combined GWAS statistics for the outcome disease/trait, enhancing the power and flexibility of the analysis. As illustrated in Figure 1(b), the second variation of int2MR can utilize the following types of summary statistics as inputs:

- IV-to-exposure statistics: Standard GWAS summary statistics for the exposure trait.
- Group-specific IV-to-outcome statistics: GWAS summary statistics for the outcome stratified by group, which could be statistics specific to the reference group, the comparison group, or both.
- Group-combined IV-to-outcome statistics: Standard GWAS summary statistics for the outcome trait derived from mixed-group samples, with a known proportion of group composition, *ρ*.

For the *j*-th SNP as an IV, we jointly model the IV-to-outcome effect for each input study and the IV-to-exposure effect as follows:

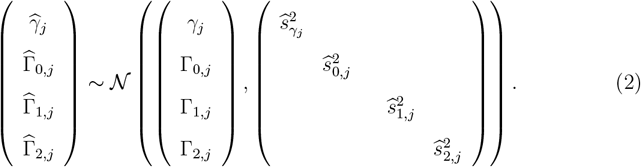

Note that the covariate matrices above may have non-zero off-diagonal elements if the input GWAS datasets have overlapping samples. These details were omitted here for clarity in presenting the main model.

The true IV-to-outcome effect in each study with varying proportions of comparison group samples is specified as:

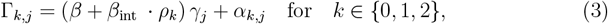

where *ρ*_*k*_ represents the proportion of comparison group samples in the *k*-th study. For the study with only reference group samples (*k* = 0), *ρ*_0_ = 0; for the study with only comparison group samples (*k* = 1), *ρ*_1_ = 1; and for the study with group-combined samples (*k* = 2), *ρ*_2_ reflects the proportion of the comparison group samples in the study. Here,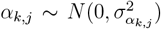is the uncorrelated pleiotropic effect for each study. The causal effect of the reference group is *β*, and the causal effect of the comparison group is *β* + *β*_int_. The interaction effect *β*_int_ captures the difference in causal effects comparing the comparison group versus the reference group.

To obtain the parameter estimates and inference of the above models, we build a Bayesian hierarchical model. We implement a No-U-Turn sampler (NUTS)^46^, a variant of the Hamiltonian Monte Carlo method ^47^, to generate posterior samples for inference. By the Bernstein–von Mises theorem, the posterior distribution asymptotically approaches a normal distribution. We estimate the standard errors of *β* and *β*_int_ using the observed Fisher information matrix, denoted as 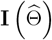, where Θ = (*β, β*_int_, *γ, α*_0_, *α*_1_, *α*_2_) represents the vector of all model parameters. The observed Fisher information matrix is computed from the Hessian of the negative posterior log-likelihood, yielding 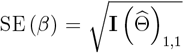 and 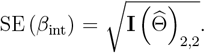. Further algorithmic details are provided in the Supplemental Information.

Model (1) integrates group-separated GWAS statistics on outcome and Model (2) integrates group-separated and combined GWAS statistics on outcome. Additionally, int2MR is generalized to integrate group-combined GWAS statistics on outcomes from two or more studies with varying group compositions. See Supplemental Information for details. For instance, when analyzing causal effects of male and female groups, we can integrate GWAS statistics on outcome from the Million Veteran Program (MVP),^48,49^ which consists of approximately 91.8% males, with another GWAS with an equal proportion of males and females. This flexibility allows int2MR to efficiently combine data from diverse sources, enhancing statistical power and enabling robust inference.

Compared to existing MR methods for detecting interaction effects, ^3,4^ a key innovation and major advantage of our model is that it requires only summary statistics. This approach provides unparalleled flexibility in assessing group-specific effects of risk factors on complex diseases, even when individual-level data on risk exposures are incomplete or unavailable. For example, in our data analysis, we evaluated agegroup-specific risk factors for Alzheimer’s disease (AD), with a focus on the oldest-old (death-age 95+). Using the ROSMAP study, we obtained age-group-separated GWAS statistics on AD for the oldest-old and those for all samples in ROSMAP as groupspecific and combined IV-to-outcome statistics, respectively. Publicly available GWAS statistics for various complex traits, sourced from other GWAS consortia, served as IV-to-exposure statistics. Notably, many of these risk exposures are not directly measured in the ROSMAP data. Yet, our method can assess their interaction effects and age-group-specific contributions to AD and pathologies.

### Allowing correlated SNPs as IVs by accounting for linkage disequilibrium (LD)

We extend the int2MR model to allow moderately correlated SNPs as IVs by modeling the LD structure:

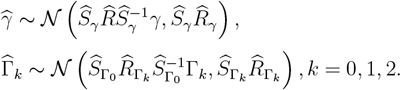

where 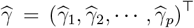 and *γ* = (*γ*_1_, *γ*_2_, · · ·, *γ*_*p*_) are the vectors of estimated and true marginal IV-to-exposure effects, respectively. 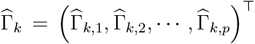 and Γ_*k*_ = (Γ_*k*,1_, Γ_*k*,2_, · · ·, Γ_*k,p*_)^?^ are the vectors of estimated and true marginal IV-tooutcome effects in the *k*-th GWAS sample (*k* ∈ {0, 1, 2}). 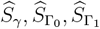 and 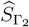 are the corresponding diagonal matrices of standard errors; and *R* is the estimated correlation matrix among all selected IVs, which could be estimated using an independent reference panel data. This framework allows for weak to moderate correlations among IVs by accounting for their correlation structures. However, the application of int2MR may not be suited when IVs are highly correlated.

## Results

### Simulations to evaluate the performance of int2MR in detecting interaction and main effects

We evaluated the performance of our proposed summary-statistics-based MR method, int2MR, by comparing it with existing methods that utilize either individual-level data or summary statistics. We simulated individual-level genotype data for GWASs of both the exposure and the outcome traits. For the outcome GWAS, we simulated GWAS datasets with group-specific effects, and generated both group-specific and group-combined GWAS summary statistics. In each simulation, we generated 300 SNPs as IVs. For the group-specific GWAS, we simulated 1,000 individuals for the comparison group and 2,000 for the reference group, and each group has group-specific effects. We calculated group-combined GWAS summary statistics by combining data from both the reference group and the comparison group. The total sample size for the mixed group was varied to investigate its effects on method performance, in particular power. In the group-combined GWAS data, individuals from the reference and comparison groups were represented in equal proportions.

From the simulated datasets, we obtained the IV-to-exposure GWAS summary statistics, as well as group-separated and group-combined GWAS summary statistics for the outcome. These summary statistics were used as inputs for our int2MR analyses. We selected the appropriate input data type, either individual-level data or summary statistics, for specific comparison method (See Supplemental Information for additional simulation details). We evaluated the type I error rates and power using a *p*-value threshold of 0.05.

Table 1(a) compares the type I error rates of int2MR using group-separated GWASs with int2MR_+20k_ integrating an additional group-combined GWAS consisting of 20,000 mixed-group samples. Additionally, we compared with the 2SLS method for interactions^26^ and standard interaction tests based on ordinary least squares (OLS) regression. We simulated various settings with and without horizontal pleiotropy and with varying strengths of confounding effects. In all settings, the proposed int2MR approach, using both group-separated and group-combined GWAS statistics for outcomes, controlled type I error rates. In contrast, the 2SLS method for interactions failed to control type I error rates in the presence of pleiotropy, and the OLS-based interaction test failed to control type I error rates in the presence of confounding. Table 1(b) compares type I error rates for testing non-zero total effects for int2MR and other existing MR methods under group-specific pleiotropy and confounding. For int2MR and int2MR+20k, the null hypothesis *H*_0_ : *β* = *β*_int_ = 0 was tested. For competing MR methods, IVW,^31^ MR-Egger, ^32^ MR-Median, ^50^ MR-RAPS,^33^ and MR-cML^36^, we tested for total effects since these methods do not assess interactions. The proposed int2MR methods consistently controlled type I error rates, whereas some competing methods showed slight inflation in type I error rates under group-specific pleiotropy or confounding.

**Table 1:** Simulation results comparing the type I error rates of different methods in different settings: with and without horizontal pleiotropy effects (*α*) and in the presence of confounders (*U*). For int2MR, we integrated group-separated GWASs for reference and comparison groups on outcome. In int2MR_+N_, the subscript “+N” represents the additional sample size from the group-combined GWAS. Among the competing methods, individual-level data-based approaches, such as 2SLS and OLS-based interaction tests, can test for interaction effects, while summary-statistics-based MR methods are limited to testing total effects. (a) Type I error rate comparisons for testing interaction effects (*β*_int_), in the presence of (group-specific) confounding and pleiotropy. (b) Type I error rate comparisons for testing total effects, in the presence of (group-specific) confounding and pleiotropy.

| Type I error rate | $\beta$ | | | $\beta_{\text{int}}$ | | |
| --- | --- | --- | --- | --- | --- | --- |
| | $\alpha = 0,$<br>$U_0 = 0,$<br>$U_1 = 0.2$ | $\alpha = 0.02,$<br>$U_0 = 0,$<br>$U_1 = 0.2$ | $\alpha = 0.02,$<br>$U_0 = 0.1,$<br>$U_1 = -0.2$ | $\alpha = 0,$<br>$U_0 = 0,$<br>$U_1 = 0.2$ | $\alpha = 0.02,$<br>$U_0 = 0,$<br>$U_1 = 0.2$ | $\alpha = 0.02,$<br>$U_0 = 0.1,$<br>$U_1 = -0.2$ |
| int2MR<sub>+20k</sub> | 0.046 | 0.050 | 0.063 | 0.048 | 0.058 | 0.063 |
| int2MR | 0.054 | 0.057 | 0.064 | 0.046 | 0.054 | 0.062 |
| 2SLS | 0.042 | 0.072 | 0.052 | 0.066 | 0.058 | 0.058 |
| OLS | 0.052 | 0.050 | 0.076 | 0.096 | 0.197 | 0.070 |
| Setting | $\alpha_0 = 0,$<br>$\alpha_1 = 0,$<br>$U = 0.2$ | $\alpha_0 = 0,$<br>$\alpha_1 = 0.02,$<br>$U = 0.2$ | $\alpha_0 = 0.02,$<br>$\alpha_1 = 0.02,$<br>$U = 0.2$ | $\alpha_0 = 0,$<br>$\alpha_1 = 0,$<br>$U = 0.2$ | $\alpha_0 = 0,$<br>$\alpha_1 = 0.02,$<br>$U = 0.2$ | $\alpha_0 = 0.02,$<br>$\alpha_1 = 0.02,$<br>$U = 0.2$ |
| int2MR<sub>+20k</sub> | 0.046 | 0.056 | 0.055 | 0.048 | 0.060 | 0.060 |
| int2MR | 0.048 | 0.045 | 0.062 | 0.048 | 0.060 | 0.062 |
| 2SLS | 0.042 | 0.080 | 0.062 | 0.072 | 0.044 | 0.080 |
| OLS | 0.174 | 0.170 | 0.170 | 0.428 | 0.400 | 0.406 |

(a)
| Type I error rate | Total Effect ($\beta + \beta_{\text{int}}$) | | | | | |
| --- | --- | --- | --- | --- | --- | --- |
| Setting | $\alpha = 0,$<br>$U_0 = 0,$<br>$U_1 = 0.2$ | $\alpha = 0.02,$<br>$U_0 = 0,$<br>$U_1 = 0.2$ | $\alpha = 0.02,$<br>$U_0 = 0.2,$<br>$U_1 = -0.2$ | $\alpha_0 = 0,$<br>$\alpha_1 = 0,$<br>$U = 0.2$ | $\alpha_0 = 0,$<br>$\alpha_1 = 0.02,$<br>$U = 0.2$ | $\alpha_0 = 0.02,$<br>$\alpha_1 = 0.02,$<br>$U = 0.2$ |
| int2MR<sub>+20k</sub> | 0.058 | 0.063 | 0.060 | 0.036 | 0.060 | 0.064 |
| int2MR | 0.048 | 0.057 | 0.062 | 0.058 | 0.053 | 0.062 |
| IVW | 0.058 | 0.072 | 0.068 | 0.054 | 0.066 | 0.084 |
| MR-Egger | 0.040 | 0.066 | 0.040 | 0.050 | 0.050 | 0.042 |
| MR-Median | 0.032 | 0.022 | 0.030 | 0.022 | 0.028 | 0.046 |
| MR-RAPS | 0.048 | 0.072 | 0.070 | 0.054 | 0.066 | 0.084 |
| MR-cML | 0.060 | 0.076 | 0.070 | 0.056 | 0.062 | 0.084 |

Figure 2(a) compares the power of different methods for detecting interaction effects. While 2SLS and OLS require individual-level data, int2MR demonstrated comparable power to the OLS-based interaction test with 2,000 samples from the reference group and 1,000 from the comparison group. When additional group-combined GWAS statistics with larger sample sizes (10k, 20k, and 50k) were integrated, int2MR showed a substantial power improvement. Figure 2(b) compares the power for detecting both main and interaction effects. For 2SLS, OLS, and int2MR, we tested non-zero main and interaction effects, while for other summary-based MR methods, we tested total effects. int2MR improved power by jointly testing for main and interaction effects.

**Figure 2:**
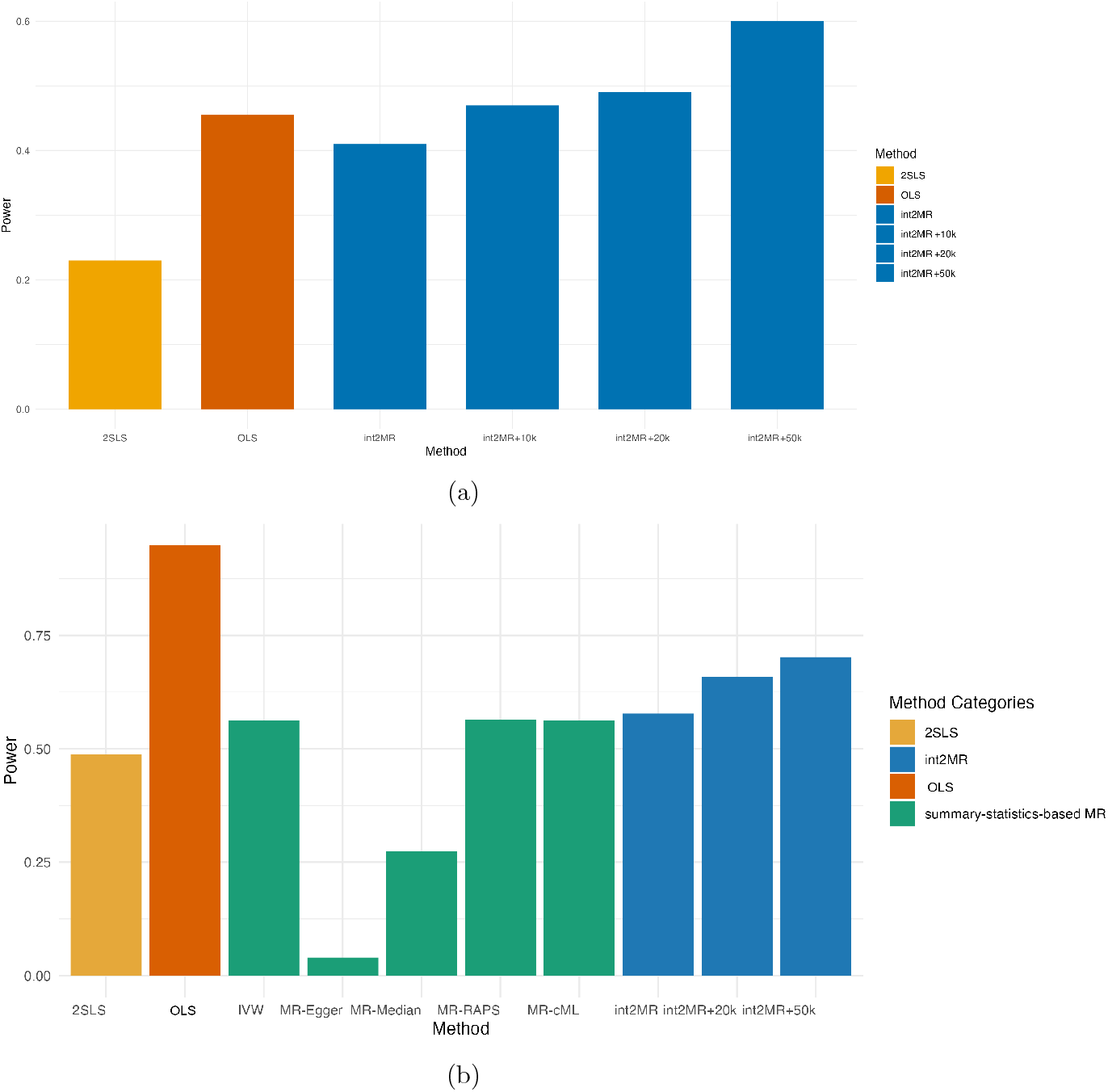
Simulation results comparing the power of int2MR with competing methods. (a) Power comparison for detecting interaction effects. The method int2MR_+N_ integrates group-separated GWAS data with a group-combined GWAS dataset, where the subscript *N* represents the sample size of the group-combined GWAS. Comparison methods are 2SLS and OLS-based interaction test, both requiring individual-level data. (b) Power comparison for detecting non-zero causal effects. We test for non-zero main or interaction effects for 2SLS, OLS-based interaction, and int2MR. For existing MR methods (colored green), we test for total effects.

In summary, the simulations showed that int2MR effectively controls type I error rates in the presence of pleiotropy and confounding. In addition, int2MR demonstrated strong power in detecting interaction effects and group-specific causal effects. The integration of group-combined and group-separated GWASs on outcomes further enhanced its power.

### Data Analysis: Identifying Risk Factors with Sex-Biased Effects on ADHD

Attention-deficit/hyperactivity disorder (ADHD) is a sex-biased condition, with males significantly more likely to be diagnosed than females. ^51–53^ These observed sex differences suggest that certain risk factors may have varying causal effects depending on sex. Identifying and understanding these sex-specific effects and their mechanisms are crucial for improving diagnosis and developing targeted interventions for both sexes. In this analysis, we leverage existing GWAS summary statistics to systematically identify potential risk factors with sex-biased effects on ADHD.

We applied the proposed int2MR method to evaluate the interaction effects between risk exposures and sex groups on ADHD. The IV-to-exposure statistics were obtained from publicly available GWASs for 33 complex traits and diseases related to immunology, metabolism, gastrointestinal health, cardiovascular health, dermatological conditions, and brain function. The IV-to-outcome GWAS statistics on ADHD were obtained from the Psychiatric Genomics Consortium (PGC), including two sex-stratified ADHD GWAS datasets as well as a sex-combined dataset. The male-only GWAS included 32,102 individuals of European ancestry, while the female-only GWAS included 21,191 individuals.^54^ Additionally, we used a larger sex-combined GWAS dataset consisting of 224,534 individuals, approximately 49.61% females^43^. We first applied int2MR using only sex-stratified ADHD GWAS statistics as the IV-to-outcome statistics. Additionally, we expanded the int2MR analysis by integrating sex-stratified GWAS statistics with a larger sex-combined GWAS to enhance the power for detecting interaction effects. For each exposure, SNPs significantly associated with the exposure (*p*-values ≤ 5 × 10^−8^) were selected as IVs, followed by LD clumping with an *r*^2^ threshold of 0.05.

At a false discovery rate (FDR) threshold of 0.1, we identified 10 traits with significant sex-biased effects on ADHD by integrating of the large sex-combined GWAS. In comparison, int2MR using only sex-stratified GWASs identified 8 of these exposures. See Table 2 for the list of ten exposures with significant sex-interaction effects on ADHD. Overall, we observed more significant *p*-values for the exposures when integrating the sex-combined GWAS. The results demonstrate improved power in detecting interaction effects by integrating sex-combined GWAS summary statistics.

**Table 2:** Results on ten risk exposures with significant sex-interaction effects on ADHD (FDR ≤ 10%) identified using int2MR. The left panel shows the results based on integrating sex-stratified with sex-combined GWAS statistics. Right shows the results based on int2MR using sex-stratified GWAS statistics only.

| Exposure | #IVs | + Sex-combined GWAS |  |  | Sex-stratified GWAS only |  |  |
| --- | --- | --- | --- | --- | --- | --- | --- |
| | | $\hat{\beta}_{\text{int}}$ | *p*-value | FDR adj. | $\hat{\beta}_{\text{int}}$ | *p*-value | FDR adj. |
| High light scatter reticulocyte count | 587 | 0.137 | 0.0006 | 0.0088 | 0.131 | 0.0010 | 0.0129 |
| Hypertension | 318 | -0.590 | 0.0007 | 0.0088 | -0.580 | 0.0008 | 0.0129 |
| Reticulocyte count | 602 | 0.117 | 0.0008 | 0.0088 | 0.116 | 0.0012 | 0.0129 |
| White blood cell count | 419 | 0.121 | 0.0014 | 0.0116 | 0.108 | 0.0059 | 0.0390 |
| Granulocyte count | 340 | 0.134 | 0.0022 | 0.0144 | 0.127 | 0.0059 | 0.0390 |
| Neutrophil + eosinophil count | 340 | 0.134 | 0.0031 | 0.0170 | 0.113 | 0.0163 | 0.0768 |
| Psoriasis | 175 | 0.204 | 0.0109 | 0.0516 | 0.186 | 0.0232 | 0.0956 |
| Myeloid white cell count | 357 | 0.112 | 0.0153 | 0.0630 | 0.112 | 0.0103 | 0.0568 |
| Basophil neutrophil count | 341 | 0.109 | 0.0231 | 0.0847 | 0.095 | 0.0541 | 0.1620 |
| Hayfever or Eczema | 290 | -0.329 | 0.0274 | 0.0903 | -0.294 | 0.0589 | 0.1620 |

**Table 3:** Risk exposures and their age-group interaction effect estimates on three AD pathologies obtained in the int2MR analysis. The three AD pathologies analyzed are Amyloid Pathology (amyloid), Global AD Pathology Burden (gpath), and Tangles Pathology (tangles). Significant exposure-by-age-group interactions were identified at a 5% FDR threshold.

| Exposure | #IVs | $\hat{\beta}_{\text{int}}$ | $p$-value | FDR adj. |
| --- | --- | --- | --- | --- |
| **Amyloid Pathology** |  |  |  |  |
| Hair or Balding Pattern 4 | 710 | -0.5482 | $6.23 \times 10^{-5}$ | 0.00299 |
| Inflammatory Bowel Disease | 243 | -0.2078 | $2.73 \times 10^{-4}$ | 0.00654 |
| Lymphocyte Count | 915 | -0.3253 | 0.00294 | 0.04706 |
| **Global AD Pathology Burden** |  |  |  |  |
| White Blood Cell Count | 833 | -0.2355 | $2.35 \times 10^{-7}$ | 0.00001 |
| Systemic Lupus Erythematosus | 251 | 0.0630 | $6.77 \times 10^{-6}$ | 0.00016 |
| Psoriasis | 315 | -0.6228 | $1.89 \times 10^{-5}$ | 0.00030 |
| Lymphocyte Count | 915 | -0.1795 | 0.00020 | 0.00157 |
| Myeloid White Cell Count | 686 | -0.2135 | 0.00018 | 0.00157 |
| Neutrophil Count | 660 | -0.2128 | 0.00014 | 0.00157 |
| Sum of Neutrophil and Eosinophil Counts | 656 | -0.2056 | 0.00825 | 0.00566 |
| Granulocyte Count | 651 | -0.2110 | 0.00182 | 0.00979 |
| Fluid Intelligence | 174 | 0.2522 | 0.00183 | 0.00979 |
| Rheumatoid Arthritis | 555 | 0.0385 | 0.00217 | 0.01042 |
| Sum of Basophil and Neutrophil Counts | 659 | -0.2138 | 0.00283 | 0.01235 |
| Sum of Eosinophil and Basophil Counts | 934 | -0.1353 | 0.05977 | 0.02391 |
| LDL Cholesterol | 95 | 0.1825 | 0.01322 | 0.04880 |
| **Tangles Pathology** |  |  |  |  |
| Chronotype | 292 | 1.3546 | 0.00020 | 0.00470 |
| Psoriasis | 315 | -1.2458 | 0.00018 | 0.00470 |
| Crohn's Disease | 192 | 0.1629 | 0.00080 | 0.01273 |

Figure 3 highlights several immune-related traits with sex-biased effects on ADHD, including white blood cell count, reticulocyte count, and granulocyte count, all of which showed larger effects (in absolute value) in males compared to females. These findings are consistent with previous reports on immune-related risk factors for ADHD, ^55 –57^ suggesting that immune system activity may play a more significant role in ADHD development among males. Prior studies have also shown that males are more prone to increased inflammatory responses, potentially driven by hormonal influences such as testosterone, which promotes a pro-inflammatory state.^58^ Additionally, hypertension has been identified as a common comorbidity in adults with ADHD. ^59^ We observed consistent effect estimates across the two int2MR analyses, with or without the integration of sex-combined GWAS data. By leveraging existing GWAS summary statistics for risk exposure traits and outcome traits from multiple data sources and studies, int2MR enables an efficient evaluation of interaction effects across a wide range of exposures. This is particularly valuable for assessing interactions of risk exposures that are not measured in the studies of outcome traits, making int2MR a powerful and versatile tool for addressing research questions that would be infeasible to study using interaction analysis methods requiring individual-level data.

**Figure 3:**
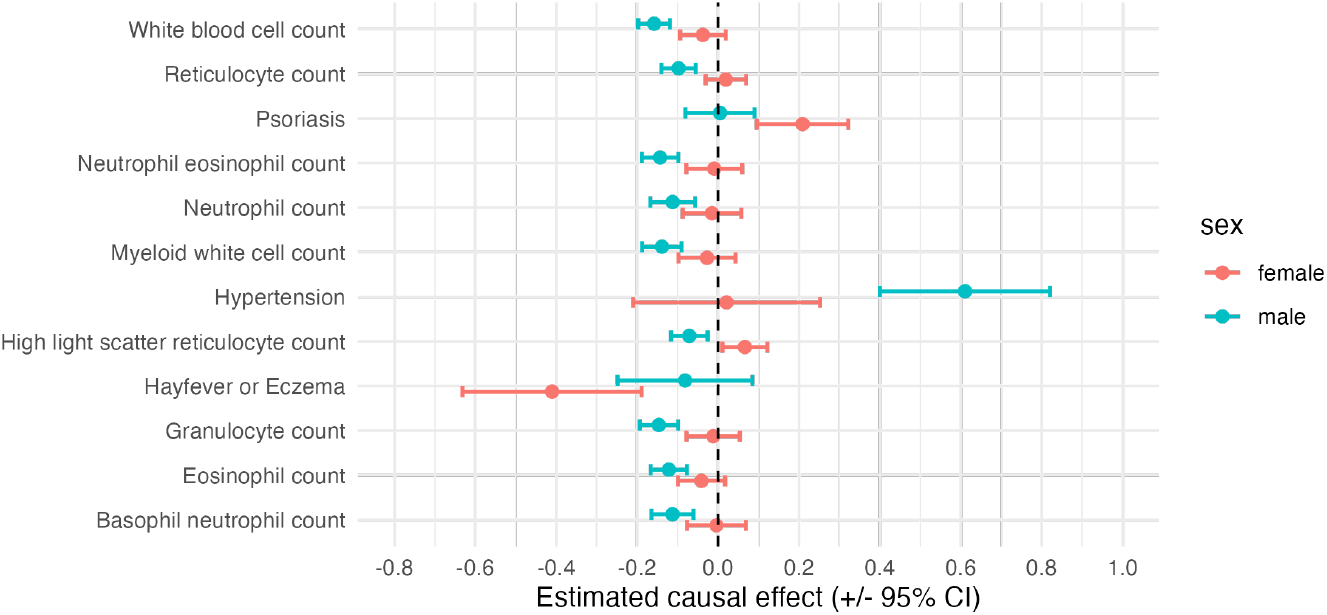
A forest plot of causal effects for risk factors with significant sex-biased effects, stratified by sex (female vs. male). Many of these risk factors are immune-related traits. Blue points represent male-specific effects, while red points represent female-specific effects.

### Data Analysis: Identifying Age-Group-Specific Risk Factors for Alzheimer’s Disease in the Oldest-Old

AD and related dementias (ADRD) present a significant public health challenge, particularly as life expectancy continues to rise. Age is the primary risk factor for ADRD. A recent analysis of data from the Religious Orders Study and Rush Memory and Aging Project (ROSMAP),^45,60,61^ involving 1,420 autopsied individuals, found that the probability of Alzheimer’s dementia and cognitive impairment increases with age. Interestingly, a nonlinear relationship was observed for AD pathology, which peaks around age 95 and then slightly declines.^62^ This pattern was evident in several AD pathology measures, including global AD pathology burden (gpath), amyloid, and PHF tau tangle density (tangle). In contrast, non-AD pathologies, except for TDP-43, continue to increase beyond age 95 in severity. Survival bias is unlikely, as the nonlinear trajectory with age is observed only for AD pathologies, while non-AD pathologies increase linearly with age. These findings highlight the need to understand the unique biology and mechanisms of neuropathologies in the oldest-old (death-age 95+) to develop effective prevention and treatment strategies for this rapidly growing age group. To identify risk factors with age-group-specific effects on AD and pathologies in the oldest-old compared to the rest, we applied the int2MR method. We examined 35 complex traits and diseases as exposures. We obtained the GWAS summary statistics for clinically diagnosed AD, three AD pathologies (gpath, amyloid, tangle), and three non-AD pathologies including TDP-43,^63–67^ hippocampal sclerosis (hspath), ^67–69^ and cortical Lewy body (dlbany),^70–72^ from the ROSMAP study. The ROSMAP GWAS data included 2,587 individuals, 408 of whom were 95 years or older at the time of death. We also obtained the GWAS summary statistics for only the oldest-old (N=408). For each exposure, SNPs with a *p*-value ≤ 10^−8^ were selected as IVs, followed by LD clumping with an *r*^2^ threshold of 0.05. The analysis was restricted to 51 exposures with at least 50 IVs after IV selection.

Figure 4 presents a heatmap showing the significance of age-group-interaction effects of selected exposures (FDR≤ 0.05) on AD pathologies (bottom panel) versus non-AD pathologies (top panel). The heatmap showed that several inflammationand immune-related traits and diseases have significant age-group-interaction effects comparing the oldest-old to the others (95+ vs. 95-). Traits such as lymphocyte and eosinophil counts showed stronger associations with AD pathologies in younger individuals (below 95). However, these associations weaken or even reverse in the oldest-old, suggesting diminished immune responses in the oldest-old. In contrast, these risk factors do not have significant age-group interaction effects on non-AD pathologies. While immune-related exposures are strongly associated with non-AD pathologies, their associations are relatively consistent across age groups (95+ vs. 95-).

**Figure 4:**
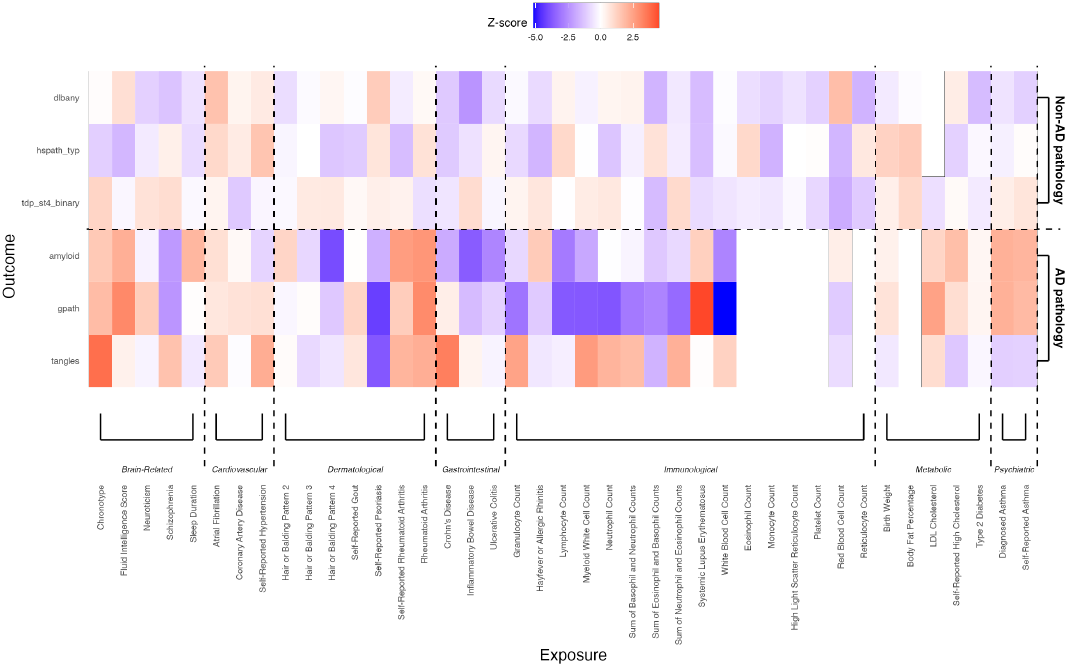
A heatmap showing age-group interaction effects on AD pathologies (bottom panel) and non-AD pathologies (top panel), comparing the oldest-old to younger age groups (95+ vs. 95-). Significant interaction effects were observed for several immune-related traits on AD pathologies. In contrast, these risk exposures showed less differences in effects comparing the two age groups, i.e., weaker age-group interactions for non-AD pathologies.

These findings underscore distinct pathological mechanisms in the oldest-old compared to the rest. The role of neuroinflammation in AD progression has been well-documented, with inflammation and immune responses driving the accumulation of AD pathology.^73–75^ However, the reduced association between immune traits and AD in the oldest-old suggests a decline in immune response and neuroinflammatory activity, potentially explaining the plateau in AD pathology accumulation in this age group. This may be due to an age-related decline in the immune system’s ability to mount an inflammatory response, along with the brain’s compensatory mechanisms.^76–81^ In contrast, non-AD pathologies continue to increase with age,^62,82^ which may result from a continuous and escalating inflammatory response that persists or intensifies with aging.^83–88^

The underlying risk factors and molecular mechanisms driving these patterns in the oldest-old remain largely unexplored. Understanding the biology of AD and related pathologies is critical for developing targeted strategies for prevention, progression, and treatment. The proposed int2MR method demonstrates its advantage by leveraging age-group-specific GWAS statistics from ROSMAP, even with a limited sample size (N=408), and integrating them with publicly available GWAS data on diverse exposures. It enables the comprehensive evaluation of age-group interaction effects across a wide range of risk exposures, many of which are not directly measured in the ROSMAP dataset. Such evaluations would be infeasible with traditional interaction tests.

## Discussion

In this study, we introduced int2MR, an integrative MR method for detecting exposureby-group interaction effects by leveraging GWAS summary statistics. int2MR combines group-separated and group-combined GWAS statistics on outcomes with GWAS statistics on exposures. The ability to use only summary statistics provides unparalleled flexibility, enabling the evaluation of group-specific effects of risk factors on complex diseases when individual-level data are incomplete or unavailable. Our simulation studies demonstrated the high power and robustness of int2MR. The method consistently controlled type I error rates in various settings. In terms of power, int2MR showed reasonable performance when using group-separated GWAS summary statistics, comparable to analyses based on individual-level data. When integrating additional group-combined GWAS data on outcomes, int2MR had substantial power improvements.

We applied int2MR to identify exposure-by-sex interaction effects for ADHD, a sex-biased disorder. The analysis revealed multiple risk factors suggesting increased inflammatory responses in males. These findings are consistent with prior studies linking inflammation and immune dysregulation to ADHD. Importantly, the integration of sex-combined GWAS data improved the power to detect interaction effects, demonstrating the method’s ability to leverage additional data to improve power. We further applied int2MR to identify age-group-specific risk factors for AD pathologies, with a focus on the oldest-old. This analysis identified multiple inflammation and immune-related risk factors, suggesting that chronic inflammation is reduced in the oldest old. This may reflect an age-related decline in the immune system’s ability to mount inflammatory responses, coupled with compensatory mechanisms in the brain. In both analyses, int2MR integrated group-separated GWAS statistics on outcomes with GWAS statistics on exposures from various sources. This capability is a major advantage over existing methods that require individual-level data, allowing for the broad assessment of interactions and group-specific effects of risk factors for complex diseases.

Despite its strengths, int2MR has several challenges that present opportunities for future research. First, the current MR model assumes that IVs are not associated with unmeasured confounders, an assumption relaxed by some recent MR methods for total effects. Future work could explore model extensions in this direction. Second, int2MR currently supports only binary group variables. Expanding the framework to interactions with continuous or multi-category variables would significantly broaden its utility. Another opportunity is the application of int2MR in transcriptome-wide MR analyses, enabling the detection of genetically regulated genes and molecular risk factors with interaction effects across various cellular contexts. int2MR is a flexible, robust, and scalable tool for detecting exposure-by-group interaction effects. Its ability to integrate diverse GWAS summary statistics opens new avenues for understanding the complex interplay between risk factors and group-specific disease mechanisms.

## Data Availability

All data produced in the present study are available upon reasonable request to the authors.

https://pgc.unc.edu/for-researchers/download-results/

https://adknowledgeportal.synapse.org/Explore/Studies/DetailsPage/StudyDetails?Study_Name=(ROSMAP)

## Data and code availability

All the summary statistics used in this paper are publicly available. They can be accessed via the corresponding references. The code for int2MR is available at https://github.com/Likeli-Ke/int2MR. Our implementation of algorithms depends on rstan (available on https://CRAN.R-project.org/package=rstan).^89^

## Acknowledgements

The research of L.S.C., B.K, J.C, and Q.C was supported by NIH 1R01GM154421 and 1U01MH139345.

## Author contributions

L.S.C. conceived the project. L.S.C. and K.X developed the methods and wrote the manuscript. K.X. developed the algorithm, and conducted the simulations. K.X. and N.M analyzed the data. All authors provided valuable suggestions for the development of the methods and the data analyses. All authors reviewed and approved the final manuscript.

## References

1. Ottman, R. (2008). Gene-environment interaction: Definitions and study designs. Nature Precedings pages 1–1. 10.1038/npre.2008.2653.1.

2. Ottman, R. & Rao, D. C. (1990). An epidemiologic approach to gene-environment interaction. Genetic epidemiology 7, 177–185. 10.1002/gepi.1370070302.

3. Zhu, X., Yang, Y., Lorincz-Comi, N., Li, G., Bentley, A. R., de Vries, P. S., Brown, M., Morrison, A. C., Rotimi, C. N., Gauderman, W. J. et al. (2024). An approach to identify gene-environment interactions and reveal new biological insight in complex traits. Nature Communications 15, 3385. 10.1038/s41467-024-47806-3.

4. Gorfine, M., Qu, C., Peters, U. & Hsu, L. (2024). Unveiling challenges in Mendelian randomization for gene–environment interaction. Genetic Epidemiology 48, 164–189. 10.1002/gepi.22552.

5. Olsson, T., Barcellos, L. F. & Alfredsson, L. (2017). Interactions between genetic, lifestyle and environmental risk factors for multiple sclerosis. Nature Reviews Neurology 13, 25–36. 10.1038/nrneurol.2016.187.

6. Perera, F. P. (1997). Environment and cancer: who are susceptible? Science 278, 1068–1073. 10.1126/science.278.5340.1068.

7. Abdul, Q. A., Yu, B. P., Chung, H. Y., Jung, H. A. & Choi, J. S. (2017). Epigenetic modifications of gene expression by lifestyle and environment. Archives of pharmacal research 40, 1219–1237. 10.1007/s12272-017-0973-3.

8. Alegría-Torres, J. A., Baccarelli, A. & Bollati, V. (2011). Epigenetics and lifestyle. Epigenomics 3, 267–277. 10.2217/epi.11.22.

9. Franks, P. W., Mesa, J.-L., Harding, A. H. & Wareham, N. J. (2007). Gene–lifestyle interaction on risk of type 2 diabetes. Nutrition, metabolism and cardiovascular diseases 17, 104–124. 10.1016/j.numecd.2006.04.001.

10. Dietrich, S., Jacobs, S., Zheng, J.-S., Meidtner, K., Schwingshackl, L. & Schulze, M. B. (2019). Gene-lifestyle interaction on risk of type 2 diabetes: A systematic review. Obesity Reviews 20, 1557–1571. 10.1111/obr.12921.

11. Kim, M. S., Shim, I., Fahed, A. C., Do, R., Park, W.-Y., Natarajan, P., Khera, V. & Won, H.-H. (2024). Association of genetic risk, lifestyle, and their interaction with obesity and obesity-related morbidities. Cell metabolism 36, 1494–1503. 10.1016/j.cmet.2024.06.004.

12. Mauvais-Jarvis, F., Merz, N. B., Barnes, P. J., Brinton, R. D., Carrero, J.-J., DeMeo, D. L., De Vries, G. J., Epperson, C. N., Govindan, R., Klein, S. L. et al. (2020). Sex and gender: modifiers of health, disease, and medicine. The Lancet 396, 565–582. 10.1016/S0140-6736(20)31561-0.

13. Lau, E. S., Paniagua, S. M., Guseh, J. S., Bhambhani, V., Zanni, M. V., Courchesne, P., Lyass, A., Larson, M. G., Levy, D. & Ho, J. E. (2019). Sex differences in circulating biomarkers of cardiovascular disease. Journal of the American College of Cardiology 74, 1543–1553. 10.1016/j.jacc.2019.06.077.

14. Westergaard, D., Moseley, P., Sørup, F. K. H., Baldi, P. & Brunak, S. (2019). Population-wide analysis of differences in disease progression patterns in men and women. Nature communications 10, 666. 10.1038/s41467-019-08475-9.

15. Van Dongen, J., Nivard, M. G., Willemsen, G., Hottenga, J.-J., Helmer, Q., Dolan, C. V., Ehli, E. A., Davies, G. E., Van Iterson, M., Breeze, C. E. et al. (2016). Genetic and environmental influences interact with age and sex in shaping the human methylome. Nature communications 7, 11115. 10.1038/ncomms11115.

16. Winkler, T. W., Justice, A. E., Graff, M., Barata, L., Feitosa, M. F., Chu, S., Czajkowski, J., Esko, T., Fall, T., Kilpeläinen, T. O. et al. (2015). The influence of age and sex on genetic associations with adult body size and shape: a large-scale genome-wide interaction study. PLoS genetics 11, e1005378. 10.1371/journal.pgen.1005378.

17. Liu, C.-T., Estrada, K., Yerges-Armstrong, L. M., Amin, N., Evangelou, E., Li, G., Minster, R. L., Carless, M. A., Kammerer, C. M., Oei, L. et al. (2012). Assessment of gene-by-sex interaction effect on bone mineral density. Journal of Bone and Mineral Research 27, 2051–2064. 10.1002/jbmr.1679.

18. Oliva, M., Muñoz-Aguirre, M., Kim-Hellmuth, S., Wucher, V., Gewirtz, A. D., Cotter, D. J., Parsana, P., Kasela, S., Balliu, B., Viñuela, A. et al. (2020). The impact of sex on gene expression across human tissues. Science 369, eaba3066. 10.1126/science.aba3066.

19. Ober, C., Loisel, D. A. & Gilad, Y. (2008). Sex-specific genetic architecture of human disease. Nature Reviews Genetics 9, 911–922. 10.1038/nrg2415.

20. Hughes, T., Adler, A., Merrill, J. T., Kelly, J. A., Kaufman, K. M., Williams, A., Langefeld, C. D., Gilkeson, G. S., Sanchez, E., Martin, J. et al. (2012). Analysis of autosomal genes reveals gene–sex interactions and higher total genetic risk in men with systemic lupus erythematosus. Annals of the rheumatic diseases 71, 694–699. 10.1136/annrheumdis-2011-200385.

21. Leng, R.-X., Wang, W., Cen, H., Zhou, M., Feng, C.-C., Zhu, Y., Yang, X.-K., Yang, M., Zhai, Y., Li, B.-Z. et al. (2012). Gene–gene and gene-sex epistatic interactions of MiR146a, IRF5, IKZF1, ETS1 and IL21 in systemic lupus erythematosus. PLoS One 7, e51090. 10.1371/journal.pone.0051090.

22. Marioni, R. E., Ritchie, S. J., Joshi, P. K., Hagenaars, S. P., Okbay, A., Fischer, K., Adams, M. J., Hill, W. D., Davies, G., Consortium, S. S. G. A. et al. (2016). Genetic variants linked to education predict longevity. Proceedings of the National Academy of Sciences 113, 13366–13371. 10.1073/pnas.1605334113.

23. Fan, Q., Verhoeven, V. J., Wojciechowski, R., Barathi, V. A., Hysi, P. G., Guggenheim, J. A., Höhn, R., Vitart, V., Khawaja, A. P., Yamashiro, K. et al. (2016). Meta-analysis of gene–environment-wide association scans accounting for education level identifies additional loci for refractive error. Nature communications 7, 11008. 10.1038/ncomms11008.

24. Conti, G. & Heckman, J. J. (2010). Understanding the early origins of the education–health gradient: A framework that can also be applied to analyze gene–environment interactions. Perspectives on Psychological Science 5, 585–605. 10.1177/17456916103835.

25. Aiken, L. S. (1991). Multiple regression: Testing and interpreting interactions. sage.

26. North, T.-L., Davies, N. M., Harrison, S., Carter, A. R., Hemani, G., Sanderson, E., Tilling, K. & Howe, L. D. (2019). Using genetic instruments to estimate interactions in mendelian randomization studies. Epidemiology 30, e33–e35. 10.1097/EDE.0000000000001096.

27. Chen, L. S., Emmert-Streib, F. & Storey, J. D. (2007). Harnessing naturally randomized transcription to infer regulatory relationships among genes. Genome Biol. 8, 1–13. 10.1186/gb-2007-8-10-r219.

28. Lawlor, D. A., Harbord, R. M., Sterne, J. A., Timpson, N. & Davey Smith, G. (2008). Mendelian randomization: using genes as instruments for making causal inferences in epidemiology. Stat. Med. 27, 1133–1163. 10.1002/sim.3034.

29. Schadt, E. E., Lamb, J., Yang, X., Zhu, J., Edwards, S., GuhaThakurta, D., Sieberts, S. K., Monks, S., Reitman, M., Zhang, C. et al. (2005). An integrative genomics approach to infer causal associations between gene expression and disease. Nat. Genet. 37, 710–717. 10.1038/ng1589.

30. Davey Smith, G. & Ebrahim, S. (2003). ‘Mendelian randomization’: can genetic epidemiology contribute to understanding environmental determinants of disease? Int. J. Epidemiol. 32, 1–22. 10.1093/ije/dyg070.

31. Burgess, S., Butterworth, A. & Thompson, S. G. (2013). Mendelian randomization analysis with multiple genetic variants using summarized data. Genet. Epidemiol. 37, 658–665. 10.1002/gepi.21758.

32. Bowden, J., Davey Smith, G. & Burgess, S. (2015). Mendelian randomization with invalid instruments: effect estimation and bias detection through Egger regression. Int. J. Epidemiol. 44, 512–525. 10.1093/ije/dyv080.

33. Zhao, Q., Wang, J., Hemani, G., Bowden, J., Small, D. S. et al. (2020). Statistical inference in two-sample summary-data Mendelian randomization using robust adjusted profile score. Annals of Statistics 48, 1742–1769. 10.1214/19-AOS1866.

34. Cheng, Q., Zhang, X., Chen, L. S. & Liu, J. (2022). Mendelian randomization accounting for complex correlated horizontal pleiotropy while elucidating shared genetic etiology. Nat. Commun. 13, 6490. 10.1038/s41467-022-34164-1.

35. Wang, J., Zhao, Q., Bowden, J., Hemani, G., Davey Smith, G., Small, D. S. & Zhang, N. R. (2021). Causal inference for heritable phenotypic risk factors using heterogeneous genetic instruments. PLoS Genet. 17, e1009575. 10.1371/journal.pgen.1009575.

36. Xue, H., Shen, X. & Pan, W. (2021). Constrained maximum likelihood-based Mendelian randomization robust to both correlated and uncorrelated pleiotropic effects. Am. J. Hum. Genet. 108, 1251–1269. 10.1016/j.ajhg.2021.05.014.

37. Morrison, J., Knoblauch, N., Marcus, J. H., Stephens, M. & He, X. (2020). Mendelian randomization accounting for correlated and uncorrelated pleiotropic effects using genome-wide summary statistics. Nat. Genet. 52, 740–747. 10.1038/s41588-020-0631-4.

38. Cheng, Q., Qiu, T., Chai, X., Sun, B., Xia, Y., Shi, X. & Liu, J. (2022). MR-Corr2: a two-sample Mendelian randomization method that accounts for correlated horizontal pleiotropy using correlated instrumental variants. Bioinformatics 38, 303–310. 10.1093/bioinformatics/btab646.

39. Bucur, I. G., Claassen, T. & Heskes, T. (2020). Inferring the direction of a causal link and estimating its effect via a Bayesian Mendelian randomization approach. Statistical Methods in Medical Research 29, 1081–1111. 10.1177/0962280219851817.

40. Zhao, J., Ming, J., Hu, X., Chen, G., Liu, J. & Yang, C. (2020). Bayesian weighted Mendelian randomization for causal inference based on summary statistics. Bioinformatics 36, 1501–1508. 10.1093/bioinformatics/btz749.

41. Grant, A. J. & Burgess, S. (2024). A Bayesian approach to Mendelian randomization using summary statistics in the univariable and multivariable settings with correlated pleiotropy. The American Journal of Human Genetics 111, 165–180. 10.1016/j.ajhg.2023.12.002.

42. Berzuini, C., Guo, H., Burgess, S. & Bernardinelli, L. (2020). A Bayesian approach to Mendelian randomization with multiple pleiotropic variants. Biostatistics 21, 86–101. 10.1093/biostatistics/kxy027.

43. Demontis, D., Walters, G. B., Athanasiadis, G., Walters, R., Therrien, K., Nielsen, T. T., Farajzadeh, L., Voloudakis, G., Bendl, J., Zeng, B. et al. (2023). Genome-wide analyses of ADHD identify 27 risk loci, refine the genetic architecture and implicate several cognitive domains. Nature genetics 55, 198–208. 10.1038/s41588-023-01350-w.

44. Demontis, D., Walters, R. K., Martin, J., Mattheisen, M., Als, T. D., Agerbo, E., Baldursson, G., Belliveau, R., Bybjerg-Grauholm, J., Bækvad-Hansen, M. et al. (2019). Discovery of the first genome-wide significant risk loci for attention deficit/hyperactivity disorder. Nature genetics 51, 63–75. 10.1038/s41588-018-0269-7.

45. Bennett, D. A., Buchman, A. S., Boyle, P. A., Barnes, L. L., Wilson, R. S. & Schneider, J. A. (2018). Religious orders study and rush memory and aging project. Journal of Alzheimer’s disease 64, S161–S189. 10.3233/JAD-179939.

46. Hoffman, M. D., Gelman, A. et al. (2014). The No-U-Turn sampler: adaptively setting path lengths in Hamiltonian Monte Carlo. J. Mach. Learn. Res. 15, 1593–1623. 10.5555/2627435.2638586.

47. Betancourt, M. (2017). A conceptual introduction to Hamiltonian Monte Carlo. arXiv preprint arXiv:1701.02434 10.48550/arXiv.1701.02434.

48. Gaziano, J. M., Concato, J., Brophy, M., Fiore, L., Pyarajan, S., Breeling, J., Whitbourne, S., Deen, J., Shannon, C., Humphries, D. et al. (2016). Million Veteran Program: A mega-biobank to study genetic influences on health and disease. Journal of clinical epidemiology 70, 214–223. 10.1016/j.jclinepi.2015.09.016.

49. Verma, A., Huffman, J. E., Rodriguez, A., Conery, M., Liu, M., Ho, Y.-L., Kim, Y., Heise, D. A., Guare, L., Panickan, V. A. et al. (2024). Diversity and scale: Genetic architecture of 2068 traits in the VA Million Veteran Program. Science 385, eadj1182. 10.1016/10.1126/science.adj1182.

50. Bowden, J., Davey Smith, G., Haycock, P. C. & Burgess, S. (2016). Consistent estimation in Mendelian randomization with some invalid instruments using a weighted median estimator. Genetic epidemiology 40, 304–314. 10.1002/gepi.21965.

51. Skogli, E. W., Teicher, M. H., Andersen, P. N., Hovik, K. T. & Øie, M. (2013). ADHD in girls and boys–gender differences in co-existing symptoms and executive function measures. BMC psychiatry 13, 1–12. 10.1186/1471-244X-13-298.

52. Polanczyk, G., De Lima, M. S., Horta, B. L., Biederman, J. & Rohde, L. A. (2007). The worldwide prevalence of ADHD: a systematic review and metaregression analysis. American journal of psychiatry 164, 942–948. 10.1176/ajp.2007.164.6.942.

53. Lahey, B. B., Applegate, B., McBurnett, K., Biederman, J., Greenhill, L., Hynd, G. W., Barkley, R. A., Newcorn, J., Jensen, P. & Richters, J. (1994). DSM-IV field trials for attention deficit hyperactivity disorder in children and adolescents. The American journal of psychiatry 151, 1673–1685. 10.1176/ajp.151.11.1673.

54. Martin, J., Walters, R. K., Demontis, D., Mattheisen, M., Lee, S. H., Robinson, E., Brikell, I., Ghirardi, L., Larsson, H., Lichtenstein, P. et al. (2018). A genetic investigation of sex bias in the prevalence of attention-deficit/hyperactivity disorder. Biological psychiatry 83, 1044–1053. 10.1016/j.biopsych.2017.11.026.

55. Wang, L.-J., Yu, Y.-H., Fu, M.-L., Yeh, W.-T., Hsu, J.-L., Yang, Y.-H., Chen, W. J., Chiang, B.-L. & Pan, W.-H. (2018). Attention deficit–hyperactivity disorder is associated with allergic symptoms and low levels of hemoglobin and serotonin. Scientific reports 8, 10229. 10.1038/s41598-018-28702-5.

56. Bale, T. L. & Epperson, C. N. (2015). Sex differences and stress across the lifespan. Nature neuroscience 18, 1413–1420. 10.1038/nn.4112.

57. Bilbo, S. D. & Schwarz, J. M. (2012). The immune system and developmental programming of brain and behavior. Frontiers in neuroendocrinology 33, 267–286. https://doi.org/j.yfrne.2012.08.006.

58. Straub, R. H. (2007). The complex role of estrogens in inflammation. Endocrine reviews 28, 521–574. 10.1210/er.2007-0001.

59. Chen, Q., Hartman, C. A., Haavik, J., Harro, J., Klungsøyr, K., Hegvik, T.-A., Wanders, R., Ottosen, C., Dalsgaard, S., Faraone, S. V. et al. (2018). Common psychiatric and metabolic comorbidity of adult attention-deficit/hyperactivity disorder: a population-based cross-sectional study. PloS one 13, e0204516. 10.1038/10.1371/journal.pone.0204516.

60. A Bennett, D., A Schneider, J., Arvanitakis, Z. & S Wilson, R. (2012). Overview and findings from the religious orders study. Current Alzheimer Research 9, 628–645. 10.2174/156720512801322573.

61. A Bennett, D., A Schneider, J., S Buchman, A., L Barnes, L., A Boyle, P. & S Wilson, R. (2012). Overview and findings from the rush Memory and Aging Project. Current Alzheimer Research 9, 646–663. 10.2174/156720512801322663.

62. Farfel, J. M., Yu, L., Boyle, P. A., Leurgans, S., Shah, R. C., Schneider, J. A. & Bennett, D. A. (2019). Alzheimer’s disease frequency peaks in the tenth decade and is lower afterwards. Acta neuropathologica communications 7, 1–9. 10.1186/s40478-019-0752-0.

63. Meneses, A., Koga, S., O’Leary, J., Dickson, D. W., Bu, G. & Zhao, N. (2021). TDP-43 pathology in Alzheimer’s disease. Molecular neurodegeneration 16, 1–15. 10.1186/s13024-021-00503-x.

64. Josephs, K. A., Murray, M. E., Whitwell, J. L., Parisi, J. E., Petrucelli, L., Jack, C. R., Petersen, R. C. & Dickson, D. W. (2014). Staging TDP-43 pathology in Alzheimer’s disease. Acta neuropathologica 127, 441–450. 10.1007/s00401-013-1211-9.

65. Josephs, K. A., Whitwell, J. L., Weigand, S. D., Murray, M. E., Tosakulwong, N., Liesinger, A. M., Petrucelli, L., Senjem, M. L., Knopman, D. S., Boeve, B. F. et al. (2014). TDP-43 is a key player in the clinical features associated with Alzheimer’s disease. Acta neuropathologica 127, 811–824. 10.1007/s00401-014-1269-z.

66. McAleese, K. E., Walker, L., Erskine, D., Thomas, A. J., McKeith, I. G. & Attems, J. (2017). TDP-43 pathology in Alzheimer’s disease, dementia with Lewy bodies and ageing. Brain pathology 27, 472–479. 10.1111/bpa.12424.

67. Nag, S., Yu, L., Capuano, A. W., Wilson, R. S., Leurgans, S. E., Bennett, D. A. & Schneider, J. A. (2015). Hippocampal sclerosis and TDP-43 pathology in aging and A lzheimer disease. Annals of neurology 77, 942–952. 10.1002/ana.24388.

68. Nelson, P. T., Schmitt, F. A., Lin, Y., Abner, E. L., Jicha, G. A., Patel, E., Thomason, P. C., Neltner, J. H., Smith, C. D., Santacruz, K. S. et al. (2011). Hippocampal sclerosis in advanced age: clinical and pathological features. Brain 134, 1506–1518. 10.1093/brain/awr053.

69. Ala, T. A., Beh, G. O. & Frey, W. H. (2000). Pure hippocampal sclerosis: a rare cause of dementia mimicking Alzheimer’s disease. Neurology 54, 843–848. 10.1212/wnl.54.4.843.

70. Mattila, P., Röyttä, M., Torikka, H., Dickson, D. & Rinne, J. (1998). Cortical Lewy bodies and Alzheimer-type changes in patients with Parkinson’s disease. Acta neuropathologica 95, 576–582. 10.1007/s004010050843.

71. Kotzbauer, P. T., Trojanowski, J. Q. & Lee, V. M.-Y. (2001). Lewy body pathology in Alzheimer’s disease. Journal of Molecular Neuroscience 17, 225–232. 10.1385/jmn:17:2:225.

72. Kenny, E. R., Blamire, A. M., Firbank, M. J. & O’Brien, J. T. (2012). Functional connectivity in cortical regions in dementia with Lewy bodies and Alzheimer’s disease. Brain 135, 569–581. 10.1093/brain/awr327.

73. Heneka, M. T., Carson, M. J., El Khoury, J., Landreth, G. E., Brosseron, F., Feinstein, D. L., Jacobs, A. H., Wyss-Coray, T., Vitorica, J., Ransohoff, R. M. et al. (2015). Neuroinflammation in Alzheimer’s disease. The Lancet Neurology 14, 388–405. 10.1016/S1474-4422(15)70016-5.

74. Chen, Y. & Yu, Y. (2023). Tau and neuroinflammation in Alzheimer’s disease: Interplay mechanisms and clinical translation. Journal of neuroinflammation 20, 165. 10.1186/s12974-023-02853-3.

75. Andronie-Cioara, F. L., Ardelean, A. I., Nistor-Cseppento, C. D., Jurcau, A., Jurcau, M. C., Pascalau, N. & Marcu, F. (2023). Molecular mechanisms of neuroinflammation in aging and Alzheimer’s disease progression. International journal of molecular sciences 24, 1869. 10.3390/ijms24031869.

76. Müller, L., Di Benedetto, S. & Pawelec, G. (2019). The immune system and its dysregulation with aging. Biochemistry and cell biology of ageing: Part II clinical science pages 21–43. 10.1007/978-981-13-3681-2.

77. Santoro, A., Bientinesi, E. & Monti, D. (2021). Immunosenescence and inflammaging in the aging process: age-related diseases or longevity? Ageing research reviews 71, 101422. 10.1016/j.arr.2021.101422.

78. Smith, R., Strandberg, O., Mattsson-Carlgren, N., Leuzy, A., Palmqvist, S., Pontecorvo, M. J., Devous Sr, M. D., Ossenkoppele, R. & Hansson, O. (2020). The accumulation rate of tau aggregates is higher in females and younger amyloid-positive subjects. Brain 143, 3805–3815. 10.1093/brain/awaa327.

79. Therneau, T. M., Knopman, D. S., Lowe, V. J., Botha, H., Graff-Radford, J., Jones, D. T., Vemuri, P., Mielke, M. M., Schwarz, C. G., Senjem, M. L. et al. (2021). Relationships between β-amyloid and tau in an elderly population: An accelerated failure time model. Neuroimage 242, 118440. 10.1016/j.neuroimage.2021.118440.

80. Gao, F., Shang, S., Chen, C., Dang, L., Gao, L., Wei, S., Wang, J., Huo, K., Deng, M., Wang, J. et al. (2020). Non-linear relationship between plasma amyloid-β 40 level and cognitive decline in a cognitively normal population. Frontiers in Aging Neuroscience 12, 557005. 10.3389/fnagi.2020.557005.

81. Whitwell, J. L., Tosakulwong, N., Weigand, S. D., Graff-Radford, J., Ertekin-Taner, N., Machulda, M. M., Duffy, J. R., Schwarz, C. G., Senjem, M. L., Jack, C. R. et al. (2021). Relationship of APOE, age at onset, amyloid and clinical phenotype in Alzheimer disease. Neurobiology of aging 108, 90–98. 10.1016/j.neurobiolaging.2021.08.012.

82. Wyss-Coray, T. (2016). Ageing, neurodegeneration and brain rejuvenation. Nature 539, 180–186. 10.1038/nature20411.

83. Bi, J., Zhang, C., Lu, C., Mo, C., Zeng, J., Yao, M., Jia, B., Liu, Z., Yuan, P. & Xu, S. (2024). Age-related bone diseases: Role of inflammaging. Journal of Autoimmunity 143, 103169. 10.1016/j.jaut.2024.103169.

84. Li, X., Li, C., Zhang, W., Wang, Y., Qian, P. & Huang, H. (2023). Inflammation and aging: signaling pathways and intervention therapies. Signal Transduction and Targeted Therapy 8, 239. 10.1038/s41392-023-01502-8.

85. Amin, J., Holmes, C., Dorey, R. B., Tommasino, E., Casal, Y. R., Williams, D. M., Dupuy, C., Nicoll, J. A. & Boche, D. (2020). Neuroinflammation in dementia with Lewy bodies: a human post-mortem study. Translational psychiatry 10, 267. 10.1038/s41398-020-00954-8.

86. Wetering, J. v., Geut, H., Bol, J. J., Galis, Y., Timmermans, E., Twisk, J. W., Hepp, D. H., Morella, M. L., Pihlstrom, L., Lemstra, A. W. et al. (2024). Neuroinflammation is associated with Alzheimer’s disease co-pathology in dementia with Lewy bodies. Acta Neuropathologica Communications 12, 73. 10.1186/s40478-024-01786-z.

87. Iba, M., Kim, C., Sallin, M., Kwon, S., Verma, A., Overk, C., Rissman, R. A., Sen, R., Sen, J. M. & Masliah, E. (2020). Neuroinflammation is associated with infiltration of T cells in Lewy body disease and α-synuclein transgenic models. Journal of Neuroinflammation 17, 1–14. 10.1186/s12974-020-01888-0.

88. Sordo, L., Qian, T., Bukhari, S. A., Nguyen, K. M., Woodworth, D. C., Head, E., Kawas, C. H., Corrada, M. M., Montine, T. J. & Sajjadi, S. A. (2023). Characterization of hippocampal sclerosis of aging and its association with other neuropathologic changes and cognitive deficits in the oldest-old. Acta Neuropathologica 146, 415–432. 10.1007/s00401-023-02606-9.

89. Carpenter, B., Gelman, A., Hoffman, M. D., Lee, D., Goodrich, B., Betancourt, M., Brubaker, M. A., Guo, J., Li, P. & Riddell, A. (2017). Stan: A probabilistic programming language. Journal of statistical software 76. 10.18637/jss.v076.i01.

