## Supplemental_Information for "Integrative Mendelian Randomization for Detecting Exposure-by-group Interactions Using Group-Specific and Combined Summary Statistics"

### 1 Model Details

#### 1.1 Modeling interaction Effects: from individual-level data to GWAS summary statistics

Let $X$ represent the risk factor of interest and $Y$ denote the outcome of interest. At the individual level, we have the following structural equations

$$X \mid G, S, U = \sum_{j=1}^{p} \gamma_j G_j + \beta_{UX} \cdot U + \varepsilon_X \tag{S1}$$

$$Y \mid X, S, G, U = \beta X + \sum \alpha_j G_j + \beta_{UY} U + \beta_{SY} \cdot S + \beta_{\text{int}} \cdot X \circ S + \varepsilon_Y. \tag{S2}$$

In equations (S1) and (S2), the vector $G = (G_1, G_2, \ldots, G_p)$ contains $p$ genetic variants for an individual, and $a = (\alpha_1, \ldots, \alpha_p)$ represents the uncorrelated pleiotropy effects. Here, $S$ denotes a binary group label (taking values in $0, 1$), and $U$ is an unmeasured confounder. The parameters of interest, $\beta$ and $\beta_{\text{int}}$, capture the causal effect of exposure $X$ on outcome $Y$ and the interaction effect between $X$ and group label $S$, respectively.

In a GWAS setting with only summary statistics, we consider a scenario where a proportion $\rho$ of the reference group is represented in the GWAS samples. For example, if our aim is to estimate the female-specific causal effect in a sex-combined GWAS, $\rho$ represents the proportion of females in the population. We assume that $S$ follows a Bernoulli distribution with parameter $\rho$, independent of $G$ and $U$. Integrating out $S$ in (S2) and substituting (S1) yields the following IV-to-outcome effect model:

$$Y \mid X, G, U = \widetilde{c}_0 + \sum_{j=1}^{k} (\beta + \beta_{\text{int}} p) \gamma_j G_j + \widetilde{\beta}_U U + \widetilde{\varepsilon}_Y,$$

where $\widetilde{c}_0$ is a constant and $\widetilde{\varepsilon}_Y$ is a noise term with mean zero. This necessitates the modeling of intercept in the two-stage linear regression in our model. In a GWAS that regresses the outcome on SNPs, the intercept does not affect the estimate of the IV-to-outcome effect, $\widehat{\Gamma}_j$.

Consequently, this model yields the following causal relationship:

$$\Gamma_j = (\beta + \beta_{\text{int}} \cdot p)\, \gamma_j + \alpha_j$$

for any $j = 1, 2, \cdots, p$, where $\Gamma_j$ is the $j$-th IV-to-outcome effect, and $\gamma_j$ is the $j$-th IV-to-exposure effect. We denote the estimated IV-to-outcome and IV-to-exposure effects as $\widehat{\Gamma}_j$ and $\widehat{\gamma}_j$, respectively. Additionally, we assume that the uncorrelated pleiotropy (UHP) effect $\alpha_j$ is balanced, with $\mathbb{E}(\alpha_j) = 0$.

#### 1.2 Comparison with competing methods

We conducted a comprehensive comparison of our method against existing approaches, categorized into two primary groups: methods utilizing individual-level data and those based on summary statistics for Mendelian Randomization (MR).

Methods leveraging individual-level data allow for direct modeling of covariates and interactions. Our methods incorporate two-stage least squares (2SLS)[1] as a competing approach. When individual-level data is available, 2SLS can account for group characteristics, such as sex and age, by including them as covariates[2]. In the first stage, we predict the effect of the instrumental variable (IV) on the exposure and the effect of the IV on sex. In the second stage, we regress the effect of the IV on the outcome using the predicted IV-to-exposure effect and the predicted IV-to-gender effect, including their interaction term.

Furthermore, we compared this with a linear regression (OLS) of the outcome on exposure using individual-level data that includes a group-covariate interaction term which refers to interaction analysis in the main text. In this linear regression, we include exposure, group and exposure-group interaction as explanatory variables, with the outcome serving as a dependent variable.

Summary-statistics-based MR methods are widely used because of their computational efficiency and reduced data-sharing requirements. In the main analysis, we compared the performance of on estimating the main effect. However, these methods are generally unable to detect or infer exposure-covariate interaction effects, a critical limitation in many epidemiological applications. Our method uniquely addresses this gap by enabling the identification and inference of interaction effects using summary data, a capability that sets it apart from existing MR methods for total effects.

### 2 Supplementary Information of Simulation

We began by generating summary statistics using simulated individual-level data for a simulation study that incorporated one GWAS sample of combined groups and two GWAS samples of single group only. Specifically, genotype matrices $G_X \in \mathbb{R}^{n_X \times p}$, $G_{Y_0} \in \mathbb{R}^{n_{Y_0} \times p}$, $G_{Y_1} \in \mathbb{R}^{n_{Y_1} \times p}$, and $G_{Y_2} \in \mathbb{R}^{n_{Y_2} \times p}$ were simulated to represent the genotypes for the exposure and outcomes in the combined and single GWAS studies for Groups 0,1 , and 2, respectively. Here, $n_X$ denotes the sample size of the IV-toexposure GWAS, and $n_{Y_k}$ for $k = 0, 1, 2$ represents the sample size of the IV-to-outcome GWAS for Group $k$.

Genotype matrices were generated by categorizing continuous genotype data into dosage values $\{0, 1, 2\}$ based on minor allele frequencies (MAF) uniformly distributed in Unif $[0.1, 0.3]$. For each individual in the combined GWAS study, a label vector $S$ was simulated from a Bernoulli distribution $B(\rho)$ to indicate the presence of an interaction term, where $\rho$ is the probability of the interaction label. For each SNP $j = 1, \ldots, p$, the effect sizes $\gamma_j$ were drawn from a uniform distribution on $(-0.2, 0.1) \cup (0.1, 0.2)$, following settings established in the existing literature[3].

We then employed the following structural model to generate individual-level data:

$$X = G_X \gamma + U_X + \varepsilon_X, \tag{S3}$$

$$Y_0 = \beta \cdot X + G_{Y_0}\alpha + U_{Y_0} + \varepsilon_{Y_0}, \tag{S4}$$

$$Y_1 = (\beta + \beta_{\text{int}}) \cdot X + a_1 \cdot G_{Y_1} + U_{Y_1} + \varepsilon_{Y_1}, \tag{S5}$$

$$Y_2 = \beta \cdot X + \beta_{\text{int}} \cdot X \circ S + a_2 \cdot G_{Y_2} + \varepsilon_{Y_2}, \tag{S6}$$

where $U_X \in R^{n_x}$ and $U_Y \in R^{n_Y}$ are the vectors for the confounding effect in the samples from IV-to-exposure and IV-to-outcome, respectively. In particular, $U_{Y_0}$ and $U_{Y_1}$ are assumed to be normal vectors with i.i.d. entries of standard deviation set to be $U_0$ and $U_1$, respectively. $X$ is the exposure. $\epsilon_X \in R^{n_X \times 1}$ is the random errors. $\beta$ is the causal effect of interest shared across two groups and $\beta_{\text{int}}$ is the interaction causal effect of interest. $S$ is a data vector in $\{0,1\}^{n_{Y_2}}$ with entries sampled from a Bernoulli distribution $B(\rho)$ independently.

Furthermore, we assumed that the uncorrelated pleiotropy effects $\alpha_k$ for the combined GWAS samples in Group $k$ are dense and follow an independent standard normal distribution. The parameters $a_0$ and $a_1$ modulate the magnitude of these uncorrelated pleiotropy effects. The notation for $U_0$ and $U_1$, which control the magnitude of confounding effects, as well as $a_0$ and $a_1$, remains consistent with the notation used in the simulation results presented in the main text. For simplicity, we assume that $a_2 = a_1$.

Finally, we conducted a single-variant analysis to derive summary statistics $\left\{\widehat{\gamma}_j, \widehat{s}_{\gamma_j}\right\}$ for the exposure and $\left\{\widehat{\Gamma}_j, \widehat{s}_{\Gamma_j}\right\}$ for the outcome for each SNP $j = 1, \ldots, p$. Under our simulation setting, we set the sample size of IV-to-exposure GWAS $n_X$ to be 50000, the sample size of Group 0 $n_{Y_0}$ to be 2000, the sample size of Group 1 $n_{Y_1}$ to be 500, corresponding to the scenario that sample size from the group of interest is smaller than the reference group. The sample size of combined Group $n_{Y_2}$ may vary. In the power simulation, we set the true main causal effect $\beta = 0.1$ and the true interaction effect $\beta = -0.05$. These summary statistics serve as the input for our proposed method.

### 3 Algorithmic Details

This section outlines the Bayesian hierarchical model implemented in the int2MR method and describes the No-U-Turn Sampler (NUTS) used for efficient parameter estimation and inference. We begin by introducing a generalized MR model, which accommodates an arbitrary number of GWAS datasets and allows for flexible correlation structures ($\rho$).

#### 3.1 Bayesian Hierarchical Model with Independent SNPs

The proposed method generalizes to scenarios involving $K$ GWAS datasets for instrument-to-outcome associations. For the $j$-th SNP, the model is expressed as:

$$\begin{pmatrix} \widehat{\Gamma}_{1,j} \\ \widehat{\Gamma}_{2,j} \\ \vdots \\ \widehat{\Gamma}_{K,j} \\ \widehat{\gamma}_j \end{pmatrix} \sim \mathcal{N}\left( \begin{pmatrix} \Gamma_{1,j} \\ \Gamma_{2,j} \\ \vdots \\ \Gamma_{K,j} \\ \gamma_j \end{pmatrix}, \operatorname{diag}\left(\widehat{s}^2_{1,j}, \ldots, \widehat{s}^2_{K,j}, \widehat{s}^2_j\right) \right),$$

where $j = 1, 2, \ldots, p$. The latent effects $\Gamma_{k,j}$, $\Gamma_{0,j}$, and $\gamma_j$ satisfy the structural equation:

$$\Gamma_{k,j} = (\beta + \rho_k \cdot \beta_{\text{int}}) \cdot \gamma_j + \alpha_{k,j}, \quad k = 1, 2, \ldots, K.$$

Here, $\gamma_j$ represents the true instrument-exposure effect, while $\Gamma_{k,j}$ represents the true instrument-outcome effects for group-specific GWAS datasets. To capture uncorrelated pleiotropy (UHP), the SNP-specific pleiotropic effects, $\alpha_{k,j}$, are modeled as:

$$\alpha_{k,j} \sim \mathcal{N}(0, \sigma^2_{\alpha_k}).$$

#### 3.2 Posterior Likelihood Specification

The full-data posterior likelihood is denoted as:

$$L\left(\Theta \mid \widehat{\gamma}, \widehat{s}_{\gamma}, \widehat{\Gamma}_k, \widehat{s}_{\Gamma_k}\right),$$

where the parameters are:

$$\Theta = \left(\beta, \beta_{\text{int}}, \{\gamma_j\}_{j=1}^{p}, \{\alpha_{k,j}\}_{k=1,\ldots,K; j=1,\ldots,p}\right).$$

Assuming independence across SNPs, the likelihood decomposes as:

$$L(\Theta) \propto \prod_{k=1}^{K}\prod_{j=1}^{p} \mathbf{p}\left(\widehat{\Gamma}_{k,j} \mid \Gamma_{k,j}, \widehat{s}_{\Gamma_{k,j}}\right) \cdot \mathbf{p}\left(\widehat{\gamma}_j \mid \gamma_j, \widehat{s}_{\gamma_j}\right) \cdot \mathbf{p}\left(\Gamma_{k,j} \mid \gamma_j, \alpha_{k,j}; \beta, \beta_{\text{int}}\right) \cdot \pi(\Theta).$$

Here, $\pi(\Theta)$ incorporates the priors on all parameters and hyperparameters, including inverse-Gamma priors for variance terms and normal priors for latent effects. For instance:

$$\sigma_{\alpha_k}^2 \sim \text{InvGamma}(\alpha_{\alpha_k}, \beta_{\alpha_k}).$$

#### 3.3 Choice of Priors and Hyperparameters

The variance parameters $\sigma_{\alpha_k}^2$ are modeled using inverse-Gamma priors:

$$\sigma_{\alpha_k}^2 \sim \text{InvGamma}(\alpha_{\alpha_k}, \beta_{\alpha_k}),$$

with hyperparameters $\alpha_{\alpha_k} = \beta_{\alpha_k} = 10^{-3}$ to ensure non-informative priors, as recommended by Gelman[4].

For the latent effect $\gamma_j$, we assume that $\gamma_j \sim \mathcal{N}(0, \sigma_\gamma^2)$. Here, the prior variance $\sigma_\gamma^2$ is the variance of $\hat{\gamma}$ across all SNPs.

Furthermore, we assign uniform priors to the causal effect parameters

$$\pi(\beta), \pi(\beta_{\text{int}}) \propto 1.$$

This choice represents a non-informative approach, ensuring that the inference is driven solely by the observed data.

#### 3.4 Implementation Details

Efficient posterior sampling is critical in high-dimensional MR analyses. Traditional MCMC methods like Gibbs sampling[5] often face challenges such as slow convergence and poor mixing. To overcome these limitations, we use the No-U-Turn Sampler (NUTS), an adaptive extension of Hamiltonian Monte Carlo (HMC). NUTS dynamically tunes the trajectory length, reducing the need for manual tuning and enhancing computational efficiency in high-dimensional parameter spaces. We refer readers to The No-U-Turn Sampler paper for technical details.[6]

In practice, we take 10,000 samples from the posterior distribution (with a burn-in of 10,000 iterations) from 2 chains. We replicated the sample procedures 500 times to obtain the type I error rate and power presented in the main text.

### References

[1] Basmann, R. L. (1957). A generalized classical method of linear estimation of coefficients in a structural equation. Econometrica: Journal of the Econometric Society pages 77–83. https://doi.org/10.2307/1907743.

[2] Zhu, X., Yang, Y., Lorincz-Comi, N., Li, G., Bentley, A. R., de Vries, P. S., Brown, M., Morrison, A. C., Rotimi, C. N., Gauderman, W. J. et al. (2024). An approach to identify gene-environment interactions and reveal new biological insight in complex traits. Nature Communications *15*, 3385. https://doi.org/10.1038/s41467-024-47806-3.

[3] Xue, H., Shen, X. & Pan, W. (2021). Constrained maximum likelihood-based Mendelian randomization robust to both correlated and uncorrelated pleiotropic effects. Am. J. Hum. Genet. *108*, 1251–1269. https://doi.org/10.1016/j.ajhg.2021.05.014.

[4] Gelman, A. (2006). Prior distributions for variance parameters in hierarchical models (comment on article by Browne and Draper). Bayesian Analysis *1*, 515 – 534. https://doi.org/10.1214/06-BA117A.

[5] Grant, A. J. & Burgess, S. (2024). A Bayesian approach to Mendelian randomization using summary statistics in the univariable and multivariable settings with correlated pleiotropy. The American Journal of Human Genetics *111*, 165–180. https://doi.org/10.1016/j.ajhg.2023.12.002.

[6] Hoffman, M. D., Gelman, A. et al. (2014). The No-U-Turn sampler: adaptively setting path lengths in Hamiltonian Monte Carlo. J. Mach. Learn. Res. *15*, 1593–1623. https://doi.org/10.5555/2627435.2638586.
